# Clinical characteristics and antibiotic resistance analysis of bloodstream infection caused by Klebsiella pneumoniae in the north of Henan province

**DOI:** 10.1101/2021.06.15.21258957

**Authors:** Yanchao Wang, Shanmei Wang, Wenjuan Yan, Youhua Yuan, Junzheng Yang

## Abstract

**Objectives:** To analyze clinical characteristics of bloodstream infection caused by Klebsiella pneumoniae and antibiotic resistance of Klebsiella pneumoniae in the north of Henan province, provide the basis for rational selection of antimicrobial drugs.

**Methods:** Klebsiella pneumoniae was isolated from 195 patients with bloodstream infection caused by Klebsiella pneumoniae in 2017 in the north of Henan Province, Phoenix100 blood culture and identification system were used for bacterial identification and drug sensitivity test was used for antibiotic resistance detection, and WHONET 5.6 software was used for data analysis of antibiotic resistance and antibiotic sensitivity; the medical history of patients, antibiotic use and laboratory examination results of 195 cases of patients with bloodstream infection caused by Klebsiella pneumoniae were also retrospectively analyzed.

**Results:** The patients with bloodstream infection caused by Klebsiella pneumoniae were mainly distributed in ICU, surgical department and Internal medicine department. There were 110 patients with bloodstream infection caused by Klebsiella pneumoniae accompanied with underlying diseases, accounting for 56.41% in 195 patients with bloodstream infection caused by Klebsiella pneumoniae, and 87 (87/110, 77.3%) patients accompanied with hypertension and diabetes. Drug sensitivity test showed that in 195 patients with bloodstream infection caused by Klebsiella pneumoniae, the top three antibiotics of the drug resistance rate of Klebsiella pneumoniae were cefazolin (74%), amoxicillin/clavulanic acid (70.1%), ampicillin /sulbactam (68.5%); the lower three antibiotics of drug resistance rate were imipenem (52%), cefepime (53%) and amikacin (33%); there were 81 strains of Klebsiella pneumoniae produced ESBLs, accounting for 41.53% in 195 strains of Klebsiella pneumoniae. The drug resistance rate of ESBLs positive strains was significantly higher than that of ESBL negative strains. It should be pointed out that the resistance of ESBLs positive strains of Klebsiella pneumoniae to cefazole reached 100%, followed by gentamicin (71.4%), ciprofloxacin (70.4%) and levofloxacin (69.1%); The resistance of ESBLs negative strains to cefazole was 18.2%, followed by gentamicin (3.5%), ciprofloxacin (3.0%) and levofloxacin (19.3%).

**Conclusions:** The number of total bacteria isolated from departments with large number of patients is relatively large, and the number of pneumonia patients caused by Klebsiella is also increased; ESBLs positive strains in this hospital are still the main reasons for the drug resistance of Klebsiella pneumoniae, reducing the antibiotics use of cefazol, gentamycin, ciprofloxacin and levofloxacin can effectively reduce the resistance of Klebsiella pneumoniae in the hospital. At the same time, we should pay attention to some contraindications for treating hypertension, diabetes drugs and antibiotics; the clinical staff should pay attention to the timely blood culture test, rational drug use can reduce the emergence of drug-resistant strains and prevent the outbreak of nosocomial infection.

## 1. Introduction

Klebsiella pneumoniae is one of the most common gram-negative bacteria causing clinical infection, bloodstream infection is one of the most serious manifestations of infectious diseases. The recent reports demonstrated that bloodstream infection caused by Klebsiella pneumoniae ranks the second in hospital gram-negative bacteremia, and the rising resistance rate of carbapenems makes the clinical anti-infection treatment more and more difficult^[1–3]^. In order to understand the current situation of drug resistance and guide rational drug use in the north of Henan province, we analyzed the clinical characteristics and bacterial resistance of 195 patients with bloodstream infection caused by Klebsiella pneumoniae in our hospital in 2017, and the reports are as follows.

## 2 Materials and methods

### 1.1 Research subjects

195 cases of patients with bloodstream infection caused by Klebsiella pneumoniae in our hospital in 2017.

### 1.2 Methods

The clinical diagnosis, department distribution, basic diseases, use of antibiotics and drug resistance of all patients with Klebsiella pneumoniae in our hospital in 2017 were analyzed; the Phoenix 100 automatic identification system was used to identify and measure bacteria strains, and drug sensitivity system (BD company) was used for antibiotic resistance detection; the ESBLs production and drug resistance were analyzed according to the experimental data. WHONET 5.6 software was used for drug sensitivity data analysis. The quality control strains were Escherichia coli ATCC25922 and Pseudomonas aeruginosa 27853.

## 3 Results

### 2.1 Genaral informatiom

In 2017, there were 1113 strains of bacteria were isolated from patients, and 195 strains were Klebsiella pneumoniae, accounting for 17.52% in total isolated bacteria; in 195 patients with bloodstream infection caused by Klebsiella pneumoniae, there were 134 males and 61 females, aged from 9 to 93 years old, the average age were 55 years old.

### 2.2 Wards distribution

In 195 patients, there were 88 cases (16.1%) in ICU department, 42 cases (14.3%) in surgery department, 50 cases (20.8%) in internal medicine department, 10 cases (62.5%) in infection department and 5 cases (27.8%) in oncology department (Table 1); especially, the highest proportion of Klebsiella pneumoniae was 62.5% in infection department in this hospital.

**Table 1.** Department Distribution of 195 patients with bloodstream infection caused by Klebsiella pneumoniae

| Department | Total number of<br>bacteria strains | Number of *Klebsiella*<br>*pneumoniae* strains | Proportion (%) |
| --- | --- | --- | --- |
| ICU | 545 | 88 | 16.15 |
| Surgery department | 294 | 42 | 14.29 |
| Internal medicine<br>department | 240 | 50 | 20.83 |
| Infectious disease<br>department | 16 | 10 | 62.50 |
| Oncology<br>department | 18 | 5 | 27.77 |
| Total | 1113 | 195 |  |

### 2.3 Accompanied diseases

Among 195 patients with bloodstream infection caused by Klebsiella pneumoniae, 110 patients had accompanied basic diseases. There were 46 cases patients accompanied with hypertension (43.6%); 21 cases patients accompanied with diabetes(19.1%); 13 cases patients accompanied with digestive system diseases (11.8%) (Table 2).

**Table 2.** Statistics of 195 patients accompanied with basic diseases

| Types of basic diseases | cases | Proportion (%) |
| --- | --- | --- |
| Hypertension | 46 | (41.8%) |
| Diabetes mellitus | 21 | (19.1%) |
| Hypertension and diabetes<br>mellitus | 20 | (18.2%) |
| Digestive system diseases | 13 | (11.8%) |
| Lung diseases | 3 | (2.7%) |
| Cardiovascular disease | 7 | (6.4%) |

### 2.4 Drug sensitivity results

Drug sensitivity results showed that the top three antibiotics of drug resistance rate of Klebsiella pneumoniae were cefazolin (74%), amoxicillin/clavulanic acid (70.1%), ampicillin/sulbactam (68.5%), and the lower three antibiotics of drug resistance rate of Klebsiella pneumoniae were Amikacin (33%), imipenem (52%) and cefepime (53%)(Table 3).

**Table 3.** Antimicrobial resistance of 195 strains of Klebsiella pneumoniae

| Antibiotics | Resistance(%) |
| --- | --- |
| Cefazolin | 74.0 |
| Amoxicillin/clavulanic acid | 70.1 |
| Ampicillin/sulbactam | 68.5 |
| ceftriaxone | 66.9 |
| Aztreonam | 64.6 |
| ciprofloxacin | 59.8 |
| Levofloxacin | 59.0 |
| Meropenem | 55.9 |
| gentamicin | 55.1 |
| Piperacillin/tazobactam | 53.5 |
| Cefepime | 52.8 |
| Imipenem | 52.0 |
| Amikacin | 33.1 |

### 2.5 ESBL test results

Among 195 strains of Klebsiella pneumoniae, 81 (41.5%) were ESBLs positive strains and 104 (58.5%) were ESBLs negative strains; the antibiotics resistance rate of ESBLs positive strains was significantly higher than that of ESBL negative strains. It should be pointed out that the resistance of ESBLs positive strains of Klebsiella pneumoniae to cefazole reached 100%, followed by gentamicin (71.4%), ciprofloxacin (70.4%) and levofloxacin (69.1%); The resistance of ESBLs negative strains to cefazole was 18.2%, followed by gentamicin (3.5%), ciprofloxacin (3.0%) and levofloxacin (19.3%), the data summarized at Table 4.

**Table 4.** Antibiotic resistance and antibiotic sensitivity of ESBL positive strains and ESBL negative strains in 195 strains of Klebsiella pneumoniae

| Antibiotics | ESBL positive strains |  |  | ESBL negative strains |  |  |
| --- | --- | --- | --- | --- | --- | --- |
|  | (n=81) |  |  | (n=114) |  |  |
|  | R | I | S | R | I | S |
| Piperacillin/tazobactam | 45.7 | 14.3 | 40.0 | 6.0 | 21.3 | 72.7 |
| Meropenem | 59.3 | 2.4 | 38.3 | 29.8 | 7.0 | 63.2 |
| Amoxicillin /clavulanic acid | 66.7 | 14.8 | 18.5 | 40.4 | 5.2 | 54.4 |
| Ampicillin/sulbactam | 60.0 | 5.7 | 34.3 | 15.7 | 2.5 | 81.8 |
| Cefepime | 65.4 | 2.5 | 32.1 | 59.6 | 0 | 40.4 |
| Aztreonam | 84 | 0 | 16 | 39.7 | 0.7 | 59.6 |
| Imipenem | 59.3 | 2.4 | 38.3 | 29.8 | 7.0 | 63.2 |
| Amikacin | 43.2 | 3.7 | 53.1 | 19.3 | 17.5 | 63.2 |
| gentamicin | 71.4 | 2.9 | 25.7 | 15.2 | 3.0 | 81.8 |
| ciprofloxacin | 70.4 | 7.4 | 22.2 | 39.5 | 3.5 | 57 |
| Levofloxacin | 69.1 | 1.3 | 29.6 | 42.1 | 19.3 | 38.6 |
| ceftriaxone | 65.7 | 0.0 | 34.3 | 33.3 | 0.0 | 66.7 |
| Cefazolin | 100.0 | 0.0 | 0.0 | 27.3 | 18.2 | 54.5 |

S: Susceptible, I: Intermediate, R: Resistant.

## 3 Discussion

Klebsiella pneumoniae is one of the most common gram-negative bacteria causing clinical infection. Bloodstream infection is one of the most serious manifestations of infectious diseases, while Klebsiella pneumoniae is the second most prevalent group of nosocomial gram negative bacilli, with severe illness and poor prognosis^[1–4]^. Due to the difference of patients in different departments and the actual situation of patients, the distribution of patients with bloodstream infection caused by Klebsiella pneumoniae varies with different departments. In this investigation, ICU is the Department with the largest number of total bacteria isolated from patients with bloodstream infection caused by Klebsiella pneumoniae in our hospital, Among them, Klebsiella pneumoniae accounted for 16.15% of the total bacteria in ICU departments; 29% in surgery department; 83% in internal medicine department; It is worth mentioning that although the total number strains of bacteria isolated from the infection department is 16 cases, the number strains of Klebsiella pneumoniae is 10 cases, accounting for 62.50% of the total number of bacteria in the Department; Of the 195 patients with bloodstream infection caused by Klebsiella pneumoniae, 88 (45.13%) patients were in ICU, 50 (25.64%) patients were in internal medicine department, 42 (21.53%) patients were in surgery department, 10 (5.12%) patients were in infection department and 5 (2.56%) patients were in oncology department. These results suggest that the number of total bacteria isolated from departments with large number of patients is relatively large, and the number of pneumonia patients caused by Klebsiella is correspondingly increased; in this statistics, the total number of bacteria isolated from the infection department was 16 cases, but the number of Klebsiella pneumoniae accounted for 62.50%, indicated that the infection department should pay attention to the prevention of pneumonia caused by Klebsiella pneumoniae, and the relevant medical staff should take protective measures.

The antibiotics sensitivity of Klebsiella pneumoniae in patients with bloodstream infection to various antibiotics in different areas and hospitals is different. The results showed that the top three antibiotics of drug resistance rate of Klebsiella pneumoniae were cefazolin (74%), amoxicillin/clavulanic acid (70.1%), ampicillin/sulbactam (68.5%), and the lower three antibiotics of drug resistance rate of Klebsiella pneumoniae were Amikacin (33%), imipenem (52%) and cefepime (53%).

Generation of extended spectrum β-Lactamases (ESBLs) are one of the most important mechanisms of drug resistance in Klebsiella pneumoniae. ESBLs are plasmid mediated, which can hydrolyze penicillins, cephalosporins and monocyclic amides, and can be used to treat them. A class of inhibitors of lactamases β-Lactamases. In 2015, the detection rate of ESBLs producing Klebsiella pneumoniae was 27.4%^[5, 6]^. Whether the laboratory has the ability to identify these ESBLs producing Klebsiella pneumoniae is a key step in reasonable management of patients, prevention and control of infection^[7]^. In this statistics, 81 ESBLs positive strains (41.5%) were detected, which was the same as that reported before (43.58%)^[4]^. The resistance of ESBLs positive strains to cefazole reached 100%, followed by gentamicin (71.4%), ciprofloxacin (70.4%) and levofloxacin (69.1%); The resistance of ESBLs negative strains to cefazole was 18.2%, followed by gentamicin (3.5%), ciprofloxacin (3.0%) and levofloxacin (19.3%); It is suggested that ESBLs positive strains are still the main cause of drug resistance of Klebsiella pneumoniae in our hospital. Reducing the use of cefazole, gentamicin, ciprofloxacin and levofloxacin can effectively reduce the drug resistance of Klebsiella pneumoniae. At the same time, we should pay attention to some contraindications for the treatment of hypertension, diabetes drugs and antibiotics. The results showed that 56% of the 195 cases of Klebsiella pneumoniae were accompanied by underlying diseases, mainly hypertension (46/195, 41.8%), followed by diabetes (21/195, 19.1%), hypertension and diabetes mellitus (20/195, 18.2%). Digestive system diseases (13/195, 11.8%).

The extensive use of antibiotics has made the drug resistance of bacteria more and more serious. This statistics shows that the drug resistance rate of Klebsiella pneumoniae to carbapenems such as meropenem (54%) and imipenem (52%) continues to rise^[8–9]^, which brings great difficulty to clinical treatment. Therefore, the clinical should pay attention to the timely submission of blood culture, According to the results of drug sensitivity and accompanying diseases, rational drug use can reduce the emergence of drug-resistant strains and prevent the outbreak of nosocomial infection.

## Data Availability

The data used to support the findings of this study are available from the corresponding author upon request

## 4. Acknowledgement

None.

